# Longitudinal brain structural changes during clozapine treatment: associations with neuroreceptor architecture and clinical response

**DOI:** 10.64898/2026.06.06.26354980

**Authors:** Bridget King, Dara M. Cannon, Nicolas A. Crossley, Alfonso Gonzalez-Valderrama, Brian Hallahan, Wi Hoon Jung, Matthew J. Kempton, Seoyoung Kim, Andrew J. Lawrence, James H. MacCabe, Colm McDonald, Cristián Mena, Shinichiro Nakajima, Andrew Papale, Samira Raminfard, Deepak K. Sarpal, Hyejin Sim, Giulia Tronchin, Lauri Tuominen, Euitae Kim, Alice Egerton

**Affiliations:** Department of Psychosis Studies, Institute of Psychiatry, Psychology and Neuroscience, King’s College London, London, United Kingdom; Clinical Neuroimaging Laboratory, Centre for Neuroimaging, Cognition and Genomics (NICOG), Galway; Department of Psychiatry, Pontificia, Universidad Catolica de Chile, Santiago, Chile; Centro de Interés Nacional para Investigación e Innovación en Niñez, Adolescencia, Resiliencia y Adversidad (IINARA), Santiago, Chile; School of Medicine, Finis Terrae University, Santiago, Chile; Dr José Horwitz Barack Psychiatric Institute, Santiago Chile; Department of Psychology, Gachon University, Gyeonggi-do, Republic of Korea; Department of Psychiatry, Seoul National University Bundang Hospital, Gyeonggi-do, Seoul, Republic of Korea; Department of Psychological Medicine, Institute of Psychiatry, Psychology and Neuroscience, King’s College London, London, UK; Department of Neuropsychiatry, Keio University School of Medicine, Tokyo; University of Pittsburgh School of Medicine, Pittsburgh, Pennsylvania, USA; Department of Brain & Cognitive Sciences, College of Natural Sciences, Seoul National University, Seoul, Republic of Korea; University of Ottawa Institute of Mental Health Research, Ottawa, ON, Canada; Department of Psychiatry, College of Medicine, Seoul National University, Seoul, Republic of Korea

## Abstract

In treatment-resistant schizophrenia, clozapine treatment has been associated with longitudinal reductions in subcortical volumes, ventricular enlargement, and widespread cortical thinning. However, it is unknown how these structural changes relate to clozapine’s pharmacological profile and clinical efficacy. We combined five longitudinal datasets with MRI acquired before and on average 5 months after clozapine initiation in 143 individuals to quantify brain structural changes and their association with normative maps relating to neuroreceptor architecture and physiological systems, and improvement in symptom severity. Clozapine treatment was associated with grey matter volume reductions across multiple subcortical regions (including the amygdala, hippocampus, thalamus, caudate, putamen and nucleus accumbens), increases in pallidal volume, ventricular enlargement, and widespread cortical thinning. Cortical regions showing the greatest magnitude of thinning corresponded to areas with higher normative densities of serotonergic 5-HT_1A_, 5-HT_2A_ and 5-HT_4_ receptors. Changes in subcortical volume or cortical thickness during clozapine treatment were not associated with changes in total or positive symptom severity. In addition, baseline subcortical volume, cortical thickness, or gyrification prior to starting clozapine did not predict subsequent symptom improvement. Cortical thinning may partly reflect clozapine’s activity at serotonergic receptors, which have been implicated in cortical network stabilisation and neuroplasticity, however structural remodelling during clozapine treatment may reflect a process independent from its clinical efficacy in improving core symptoms of psychosis.

## Introduction

Approximately 30% of patients with schizophrenia are considered to have treatment-resistant schizophrenia (TRS), defined by an inadequate response to at least two non-clozapine antipsychotic trials (1). Clozapine remains the most effective intervention for TRS, with ∼50% of patients showing a clinical improvement (2,3), however the neurobiological mechanisms underlying the efficacy of clozapine remain elusive (4).

Schizophrenia is associated with volumetric reductions in the hippocampus, amygdala, thalamus and accumbens, as well as lateral ventricle enlargement and widespread cortical thinning (5–8). These structural alterations may emerge before illness onset or antipsychotic treatment (9–14). In addition to illness-related processes, cumulative antipsychotic exposure has been associated with reductions in cortical, whole-brain grey matter volume and lateral ventricular enlargement (15–21). Grey-matter volume reductions and cortical thinning often occur in parallel with clinical improvement and may reflect cortical reorganisation or neuroadaptive remodelling rather than maladaptive processes (22,23).

Several longitudinal studies in TRS, although with relatively small sample sizes (n = 22–33), have reported that, despite prior antipsychotic exposure, additional structural brain changes occur following commencing clozapine. These studies have reported volumetric reductions in the caudate (24–27), putamen (25), thalamus, and hippocampus (27) and enlargement of the lateral ventricles (25,27) during clozapine treatment. Clozapine treatment also appears associated with widespread cortical thinning (25,28,29).

Clozapine has a broad and complex pharmacological profile (30), wherein one, multiple, or interacting mechanisms may contribute to the observed brain structural changes. Clozapine has high affinity for multiple serotonergic (5-HT_2A_, 5-HT_2C_, 5-HT_6_, 5-HT_7_), muscarinic (M_1_, M_4_), histaminic (H_1_) and α_1_-adrenergic receptors, acting predominantly as an antagonist or inverse agonist, with partial agonism at the M_4_ (31). It also exhibits moderate affinity as an antagonist at dopaminergic D_2_, D_3_ and D_4_, muscarinic M_2_/M_3_ and adrenergic α_2A_/α_2C_receptors as well as agonism at 5-HT_1A_ (30). Clozapine may also indirectly influence glutamatergic (30,32–35), GABAergic (36–38), metabolic (25), perfusion-related (39,40) and astrocytic processes (41).

Mapping the spatial topology of brain structural changes on to normative maps of neurochemical and physiological features can provide insights into the mechanisms underlying differential regional vulnerability (42). Using this approach, a recent cross-sectional study applying a discovery and replication cohort found that reductions in cortical thickness associated with lifetime antipsychotic exposure showed spatial correspondence with regions with high normative expression of serotonergic (5-HT_2A_ and 5-HT_4_) and nicotinic (α4β2) receptors, and in the discovery cohort only, to regional glucose metabolism (43). This would be consistent with the involvement of serotonin and acetylcholine in neuroplasticity and remodelling (44,45), and antipsychotic-induced metabolic stress (46). The discovery cohort in Tuominen et al. (43) comprised individuals at clinical high risk for psychosis or experiencing first episode psychosis, with no clozapine exposure. In the ENIGMA schizophrenia replication cohort, specific information on antipsychotic medications was unavailable (43). Thus, it is unknown whether the relationship between normative maps for serotonin and nicotinic receptor and glucose metabolism and susceptibility to antipsychotic-related cortical thinning extends to clozapine.

In addition to understanding the neurobiological mechanisms that may contribute to antipsychotic related brain structural changes, it is also important to examine potential associations with clinical response. Cross-sectional studies indicate greater reductions in subcortical volumes and cortical thickness among patients who do not respond to antipsychotic treatment (47,48). These structural differences appear to be most pronounced in TRS (49–53) and may further differentiate between clozapine responders and non-responders (54–56). Differences in cortical gyrification have also been implicated in TRS and the degree of clozapine response (57,58).

Longitudinal studies have also provided some preliminary evidence that structural trajectories may differ according to clozapine response (26,28), although these analyses have been limited by relatively small sample sizes. Greater cortical thinning in left medial frontal and right middle temporal regions has been observed in clozapine non[responders (28), and symptom improvement has been associated with reductions in caudate volume (26). However, other studies have not detected such relationships (25,49,56). Brain structural features may also predict subsequent antipsychotic response as lower baseline grey-matter volumes and cortical thickness have been linked to poor antipsychotic response and greater symptom burden (59,60), and reduced frontal/temporal gyrification predicts non-response to first-line antipsychotics (61). However, it is unclear whether structural features prior to clozapine initiation can predict subsequent clozapine response. This is important because identifying predictors of clozapine response could encourage earlier clozapine initiation in those most likely to benefit from clozapine treatment.

In this study, we combined five independent datasets to examine longitudinal brain structural changes during clozapine treatment in TRS and the neurobiological features that may contribute to clozapine-related cortical thinning. We hypothesised that clozapine treatment would be associated with reductions in subcortical volumes, enlargement of the lateral ventricles, and widespread cortical thinning. We hypothesised that the spatial pattern of cortical thinning would correspond to normative maps of neurotransmitter systems targeted by clozapine, including serotonergic, cholinergic, dopaminergic, glutamatergic and GABAergic systems. In exploratory analyses, we also examined whether cortical thinning corresponded to maps of oxygen metabolism, cerebral blood flow (CBF), translocator protein (TSPO), synaptic terminal density (SV2A) and epigenetic regulators (histone deacetylase, HDACs). We additionally hypothesised that symptom reduction during clozapine treatment would be positively associated with reductions in subcortical volumes, negatively associated with cortical thinning, and that greater structural integrity prior to commencing clozapine would be associated with a better subsequent clinical response.

## Methods

Data for this study represents part of the CLIMATE (CLozapine IMAging and Treatment Effects) collaboration. Data were shared between 5 sites: London (King’s College London), Galway (University of Galway), Santiago (Pontifical Catholic University of Chile), Pittsburgh (University of Pittsburgh) and Seoul (Seoul National University). We included TRS participants with structural scans acquired before and after clozapine initiation, resulting in a sample of 147 TRS participants (N = London: 24, Galway: 33, Pittsburgh: 23, Santiago: 33, Seoul: 34).

### Included study information

Study protocols for the London (25), Galway (27,28) and Pittsburgh (62) have been previously described in detail. All studies included participants with a diagnosis of schizophrenia or schizoaffective disorder who were considered TRS and were about to initiate clozapine as part of routine clinical care. The inclusion and exclusion criteria for each study are included in eAppendix 1. Participants at the Galway, Santiago and Seoul sites were excluded if they had ever been exposed to clozapine, whereas at the Pittsburgh and London sites participants were excluded if they been exposed to clozapine in the previous 4 weeks or 3 months, respectively.

### Clinical assessments

At four sites (London, Galway, Santiago, Seoul), symptom severity was measured using the Positive and Negative Syndrome Scale (PANSS). At the remaining site (Pittsburgh) symptom severity was assessed using the Brief Psychiatric Rating Scale (BPRS) and Scale for the Assessment of Negative Symptoms (SANS). For analysis, the BPRS and SANS scales were converted into a proxy of PANSS negative and total scores, as previously described (63,64). A PANSS positive proxy score was derived from summing scores on the following BPRS items: Conceptual Disorganisation, Grandiosity, Hostility, Suspiciousness, Hallucinations, Unusual Thought Content and Excitement.

### MRI data processing and harmonisation

MRI scans were acquired at 3 Tesla (T) at the London (General Electric), Pittsburgh (Siemens Prisma), Santiago and Seoul sites (Philips Ingenia) and at 1.5T in Galway (Siemens Symph). Raw T1-weighted scans were transferred and processed centrally at the London site using the longitudinal stream in FreeSurfer (version 7.4.1; https://surfer.nmr.mgh.harvard.edu/)(65–67). This approach creates an unbiased within-subject template from both timepoints for subsequent processing, including surface reconstruction and Desikan-Killiany atlas segmentation, improving reliability and statistical power (66). Cortical thickness were extracted using the Desikan-Killiany atlas (68 regions) (68) and subcortical volumes were obtained from FreeSurfer’s subcortical segmentation pipeline (16 regions) (69). Local gyrification index (LGI) was computed using FreeSurfer’s mris_compute_lgi tool from the longitudinally processed surfaces. Quality control followed ENIGMA consortium protocols (http://enigma.ini.usc.edu/) for cortical and subcortical measures.

To mitigate site effects due to scanner differences, ComBat harmonisation was applied using the neuroHarmonize Python package (70,71). ComBat is an empirical Bayes method used to minimise scanner variance while preserving biological variation (70,72,73). Within the ComBat model, biological variance related to age, sex, and illness duration was preserved; age and illness duration were modelled as Generalized Additive Model covariates to account for potential non-linear effects. ComBat models were first trained using the baseline data and then applied to follow-up data. ComBat was applied separately to cortical thickness, subcortical volumes and LGI measures (eFigure 1). Data in 1 participant were excluded from all analyses due to QC failure, and data in 3 participants was excluded from the harmonisation process and the final dataset due to incomplete clinical information.

### Statistical analysis

Changes in subcortical volume and cortical thickness between baseline and follow-up scans were quantified using the annualised symmetrised percentage change (SPC). This approach simplifies longitudinal data in each subject into a single statistic and is suitable for evaluating linear relationships across two timepoints while accommodating differences in follow-up intervals (66). SPC reflects the annualised rate of change, calculated as:

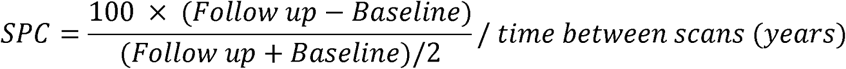

### Cortical thickness and subcortical volume changes during clozapine treatment

Statistical analyses were performed in R (version 4.3.1). Multivariate analyses of covariance (MANCOVA) first tested whether SPC values differed from zero adjusting for age and sex. Where a significant multivariate effect was observed, one-sample t-tests determined whether regional SPC values significantly deviated from zero. Regions surviving false discovery rate (FDR) correction (Benjamini-Hochberg procedure) were subsequently entered into separate MANCOVAs, covarying for age and sex.

Subsequent sensitivity analyses included exclusion of influential observations, as identified using Cook’s distance (Cook’s D > 4/n), and leave-one-site-out analyses. Additionally, as inter-scan intervals varied by site, analyses were repeated using non-annualised SPC values. Finally, to further assess robustness, linear mixed-effects models (LMMs) were fitted to the uncorrected subcortical volume and cortical thickness data, with time (baseline or follow-up), age, sex, illness duration and days between scans included as fixed effects, and random intercepts for participant and site.

### Receptor density profiles

Associations between cortical thinning and receptor density profiles were investigated using positron emission tomography (PET) maps within the neuromaps toolbox (74). We selected maps relating to: D_1_, D_2_, 5-HT_1A_, 5-HT_1B_, 5-HT_2A_, 5-HTT, 5-HT_6_, α4β2, M_1_, NMDA, mGlu5, GABA-A, CBF, glucose metabolism, oxygen metabolism, HDAC, TSPO and SV2A (eAppendix 2).

Each PET map was parcellated using the Desikan-Killiany cortical atlas (68) to align with the cortical SPC data. Pearson’s correlations between the SPC values for each cortical region and the corresponding region-wise values from each PET receptor map were computed. Spatial autocorrelation using spin-tests accounted for statistical non-independence of neighbouring data points (75). This approach generates a null distribution of correlation coefficients by randomly rotating one of the cortical maps on the spherical surface and repeating the correlation 10,000 times. The empirical correlation is then compared with this null distribution to calculate a permutation-based p-value (P_spin_) (75,76).

### Relationships between structural changes and clinical variables

Linear regression models (LRMs) were used to assess whether regional SPC in cortical thickness and subcortical volume were associated with percentage changes in positive or total symptom severity, covarying for baseline symptom scores. The percentage change in total or positive symptom severity was calculated as (follow up value − baseline value)/baseline value * 100). Minimum possible scores were subtracted prior to this calculation (77).

Sensitivity analyses excluded influential observations identified using Cook’s distance (Cook’s D > 4/n).

To examine whether clozapine dose and/or serum levels at follow-up were associated with regional SPCs, separate region-wise linear regression models were fitted with SPC values for each region entered as dependent variables and clozapine dose or serum levels included as predictors.

### Relationships between baseline structural features and subsequent changes in symptom severity

LRMs tested for relationships between baseline cortical thickness, subcortical volumes and LGI values and subsequent percentage change in total or positive symptom severity, covarying for baseline symptom scores (and ICV for subcortical volumes).

## Results

The final dataset comprised of 143 participants with TRS (mean ± SD age = 33.47 ± 10.94 years; 64.6% male, Table 1). The mean interval between scans was 152.23 ± 61.55 days, and the mean illness duration before clozapine initiation was 10.86 ± 8.92 years. Total symptom severity improved from 75.25 ± 15.42 at baseline to 58.61 ± 14.32 at follow-up (P <.001).

**Table 1.**
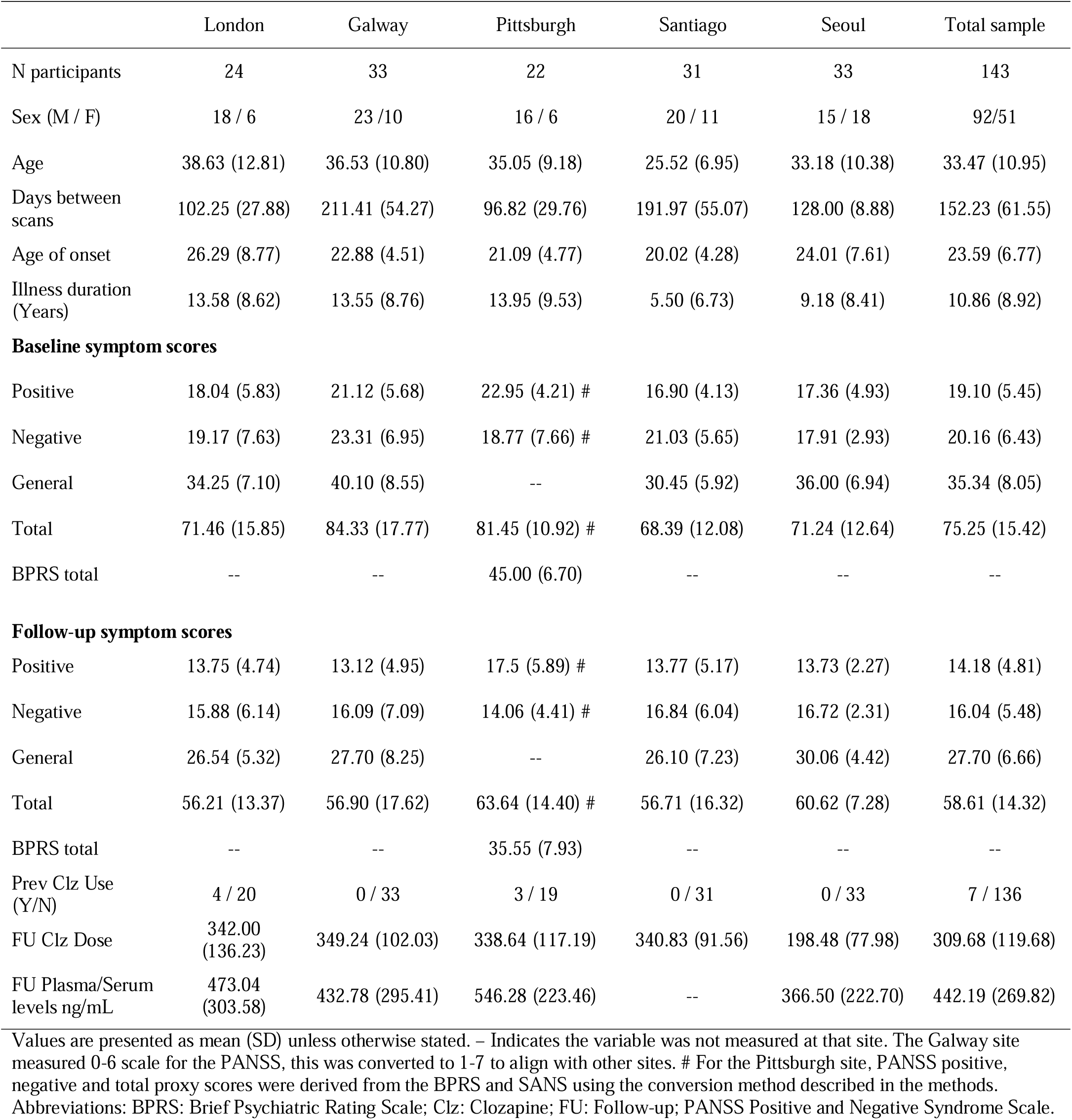
Characteristics of the sample.

|  | London | Galway | Pittsburgh | Santiago | Seoul | Total sample |
| --- | --- | --- | --- | --- | --- | --- |
| N participants | 24 | 33 | 22 | 31 | 33 | 143 |
| Sex (M / F) | 18 / 6 | 23 / 10 | 16 / 6 | 20 / 11 | 15 / 18 | 92/51 |
| Age | 38.63 (12.81) | 36.53 (10.80) | 35.05 (9.18) | 25.52 (6.95) | 33.18 (10.38) | 33.47 (10.95) |
| Days between scans | 102.25 (27.88) | 211.41 (54.27) | 96.82 (29.76) | 191.97 (55.07) | 128.00 (8.88) | 152.23 (61.55) |
| Age of onset | 26.29 (8.77) | 22.88 (4.51) | 21.09 (4.77) | 20.02 (4.28) | 24.01 (7.61) | 23.59 (6.77) |
| Illness duration (Years) | 13.58 (8.62) | 13.55 (8.76) | 13.95 (9.53) | 5.50 (6.73) | 9.18 (8.41) | 10.86 (8.92) |
| **Baseline symptom scores** |  |  |  |  |  |  |
| Positive | 18.04 (5.83) | 21.12 (5.68) | 22.95 (4.21) # | 16.90 (4.13) | 17.36 (4.93) | 19.10 (5.45) |
| Negative | 19.17 (7.63) | 23.31 (6.95) | 18.77 (7.66) # | 21.03 (5.65) | 17.91 (2.93) | 20.16 (6.43) |
| General | 34.25 (7.10) | 40.10 (8.55) | -- | 30.45 (5.92) | 36.00 (6.94) | 35.34 (8.05) |
| Total | 71.46 (15.85) | 84.33 (17.77) | 81.45 (10.92) # | 68.39 (12.08) | 71.24 (12.64) | 75.25 (15.42) |
| BPRS total | -- | -- | 45.00 (6.70) | -- | -- | -- |
| **Follow-up symptom scores** |  |  |  |  |  |  |
| Positive | 13.75 (4.74) | 13.12 (4.95) | 17.5 (5.89) # | 13.77 (5.17) | 13.73 (2.27) | 14.18 (4.81) |
| Negative | 15.88 (6.14) | 16.09 (7.09) | 14.06 (4.41) # | 16.84 (6.04) | 16.72 (2.31) | 16.04 (5.48) |
| General | 26.54 (5.32) | 27.70 (8.25) | -- | 26.10 (7.23) | 30.06 (4.42) | 27.70 (6.66) |
| Total | 56.21 (13.37) | 56.90 (17.62) | 63.64 (14.40) # | 56.71 (16.32) | 60.62 (7.28) | 58.61 (14.32) |
| BPRS total | -- | -- | 35.55 (7.93) | -- | -- | -- |
| Prev Clz Use (Y/N) | 4 / 20 | 0 / 33 | 3 / 19 | 0 / 31 | 0 / 33 | 7 / 136 |
| FU Clz Dose | 342.00 (136.23) | 349.24 (102.03) | 338.64 (117.19) | 340.83 (91.56) | 198.48 (77.98) | 309.68 (119.68) |
| FU Plasma/Serum levels ng/mL | 473.04 (303.58) | 432.78 (295.41) | 546.28 (223.46) | -- | 366.50 (222.70) | 442.19 (269.82) |

Values are presented as mean (SD) unless otherwise stated. -- Indicates the variable was not measured at that site. The Galway site measured 0-6 scale for the PANSS, this was converted to 1-7 to align with other sites. # For the Pittsburgh site, PANSS positive, negative and total proxy scores were derived from the BPRS and SANS using the conversion method described in the methods. Abbreviations: BPRS: Brief Psychiatric Rating Scale; Clz: Clozapine; FU: Follow-up; PANSS Positive and Negative Syndrome Scale.

### Changes in subcortical volumes during clozapine treatment

MANCOVA indicated significant deviation from zero across all subcortical SPC values (Pillai’s trace = 0.742, F (16, 126) = 22.64, *p* < .001). Significant changes in SPC during clozapine treatment were detected for all subcortical regions bilaterally, with small to large effect sizes. The volumes of the amygdala, hippocampus, thalamus, caudate, putamen and nucleus accumbens significantly decreased during clozapine treatment (Cohen’s *d* = -0.99 to *-* 0.23), whilst pallidum and lateral ventricle volumes significantly increased (Cohen’s *d* = 0.23 to 0.65) (Figure 1). All results remained significant after applying FDR correction, covarying age and sex (Table 2), removal of potential outliers, and when the sample was restricted to clozapine-naïve patients (n = 136; eTable 1).

**Figure 1.**
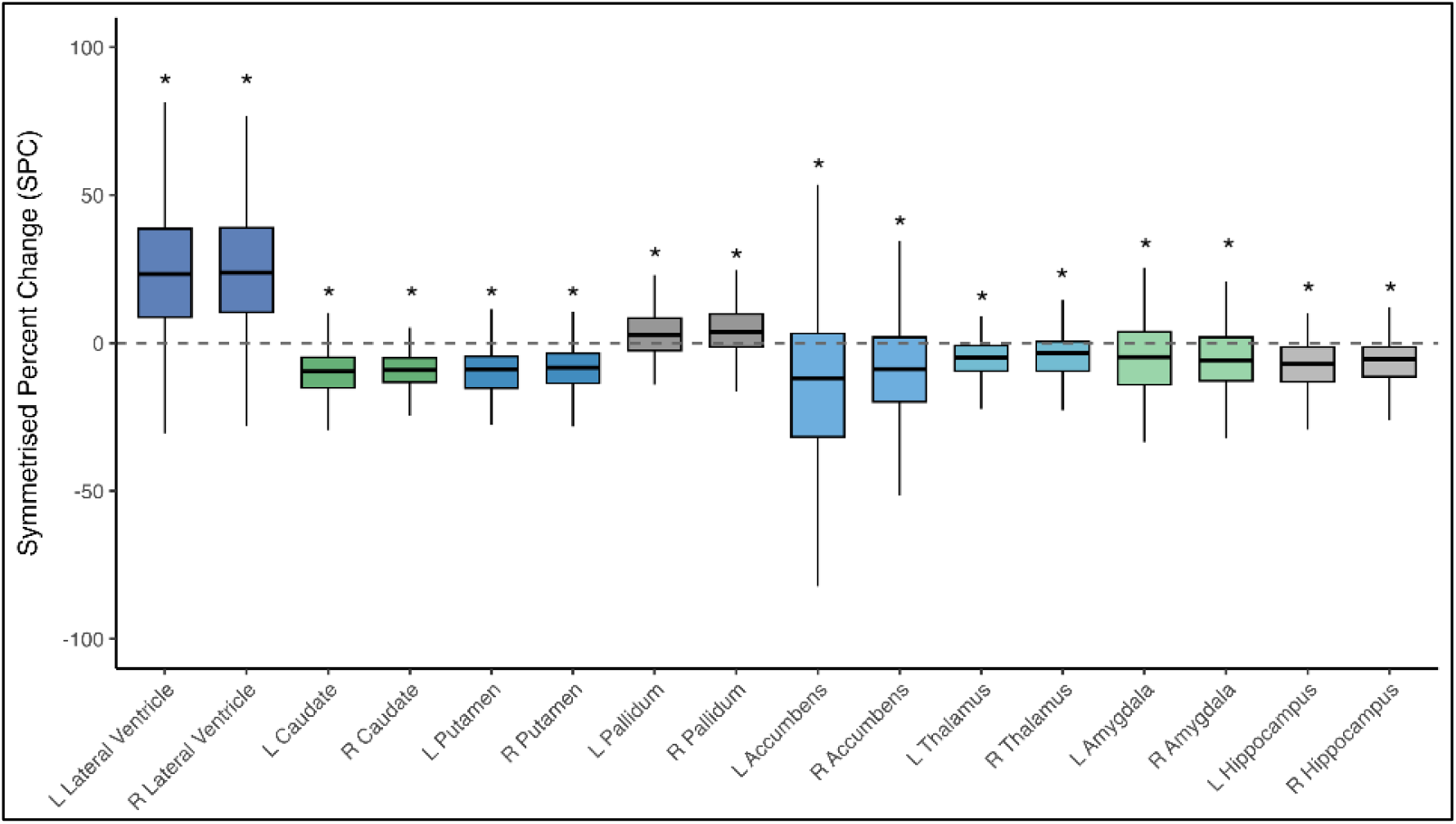
Boxplots displaying symmetrised percentage change (SPC) in subcortical regions during clozapine treatment. L=left hemisphere; R= right hemisphere. Significant SPC changes were observed across all subcortical regions (*P_FDR_<0.05). SPC was calculated as the percentage difference between follow-up and baseline values relative to their mean and annualised by dividing by the time between scans (years). Please note SPC values are 4-5 fold greater than percentage change.

**Table 2:**
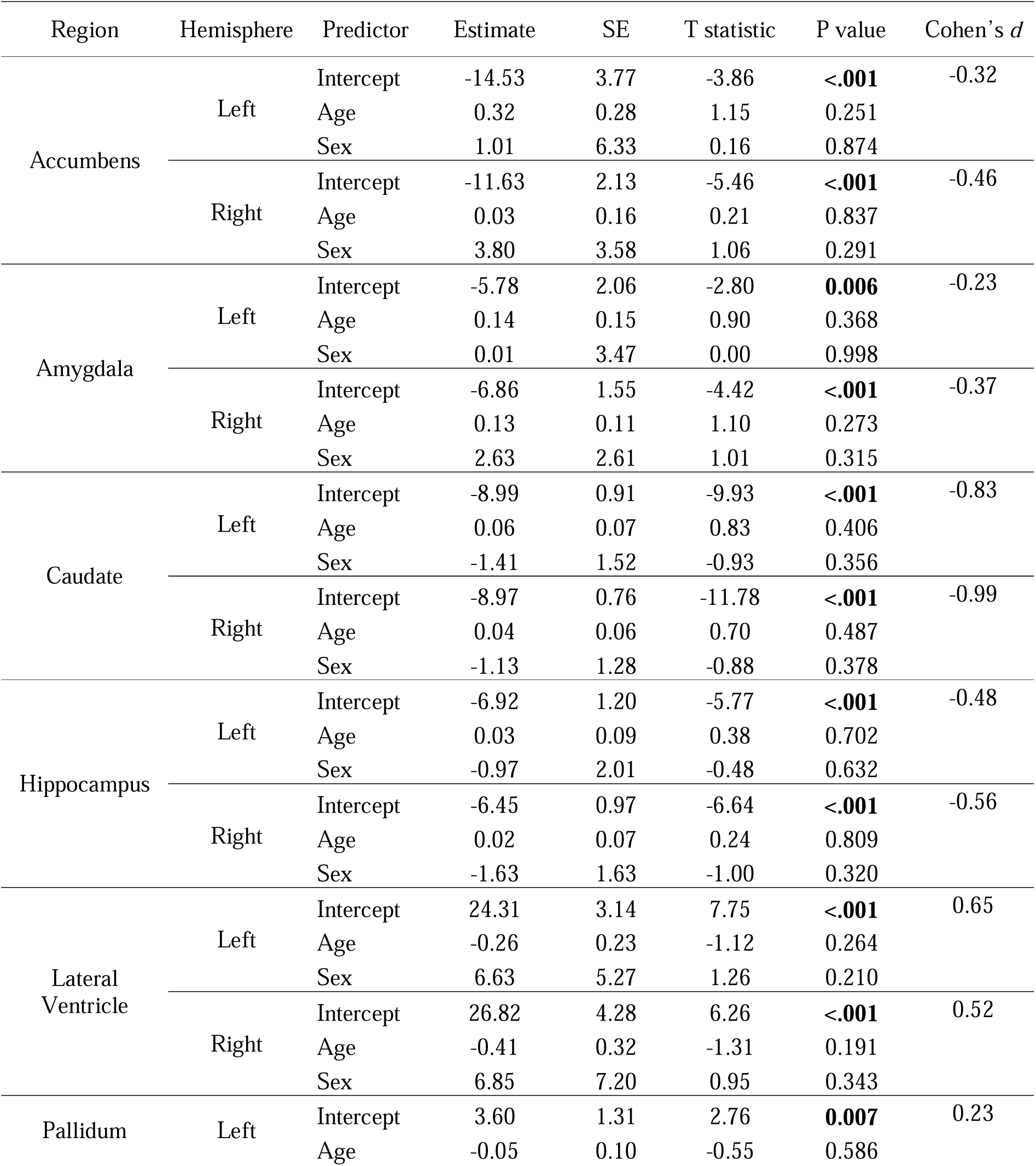
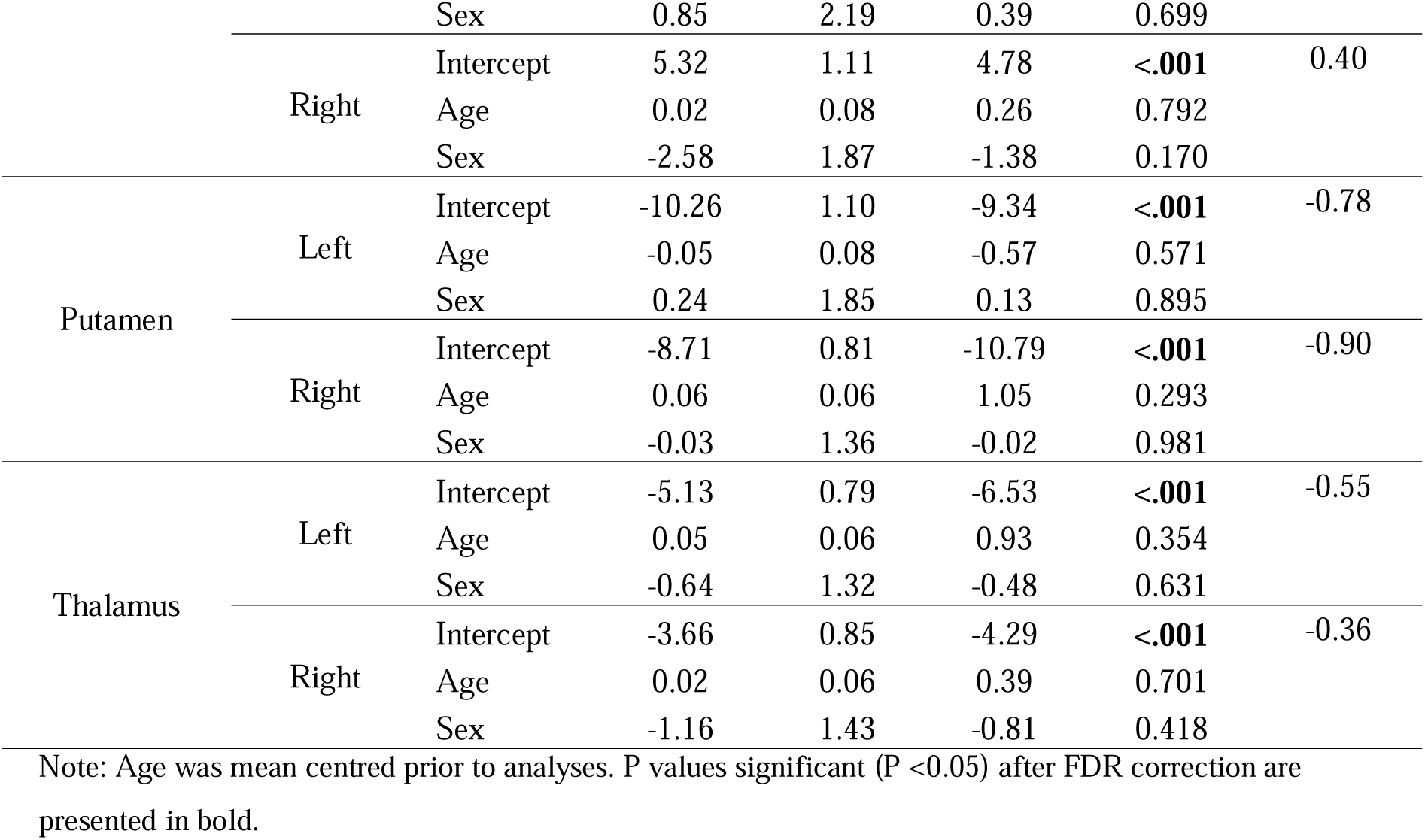
Changes in subcortical volumes during clozapine treatment.

Similar results were observed when analyses were conducted using non-annualised SPC (eTable 2). Visual inspection of non-annualised SPC plotted against days between scans showed no evidence of strong non-linear relationships that could bias annualised estimates, with effects appearing broadly stable across the timepoints analysed (eFigure 2). Furthermore, uncorrected data analysed with LMMs (eTable 3) and leave-one-site-out analyses (eTable 4) also produced similar findings.

### Cortical thinning during clozapine treatment

MANCOVA indicated significant deviation from zero across cortical thickness SPC values (Pillai’s trace = 0.792, F (67, 75) = 4.26, p = <.001). All cortical regions showed significant thinning during clozapine treatment, with small to moderate effect sizes (Cohen’s *d* = -0.68 to -0.13), except for the pericalcarine gyrus in the right hemisphere. After FDR correction, all regions remained significant except for the transverse temporal gyrus in the right hemisphere. These findings were unchanged after covarying for age and sex (Figure 2, eTable 5), removal of potential outliers, and when the sample was restricted to clozapine-naïve patients (n = 136; eTable 1).

**Figure 2.**
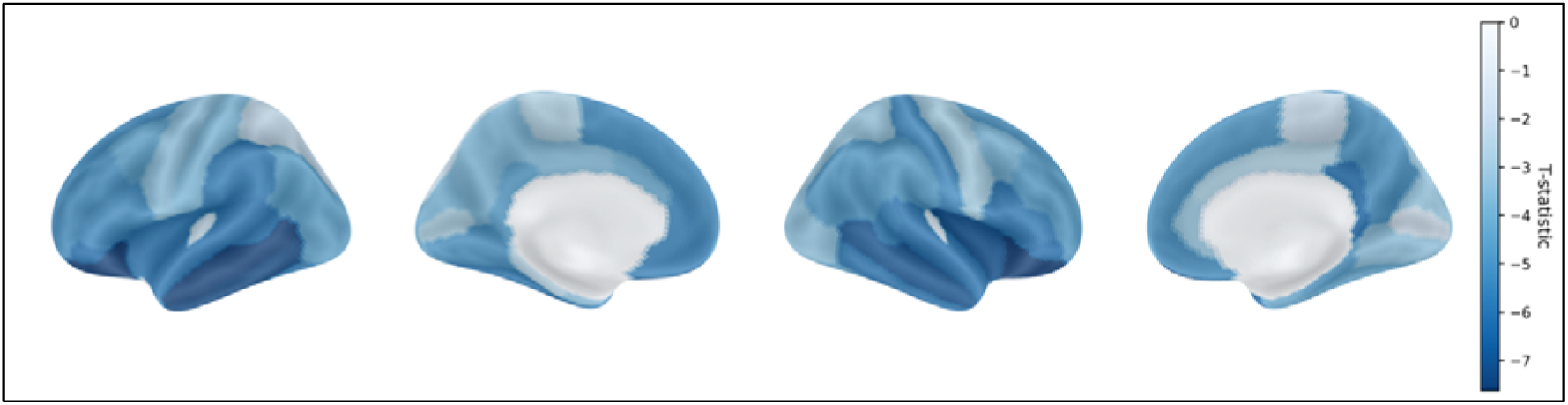
Brain render displaying cortical thinning during clozapine treatment, after covarying for age and sex. The colour intensity indicates the magnitude of the T-statistic, with darker blue indicating greater cortical thinning. Widespread cortical thinning was observed, with greatest thinning in frontal and temporal association cortices and more moderate effects in parietal and occipital regions (eTable 5).

Similar results were observed when analyses were conducted using non-annualised SPC (eTable 2), with visual inspection of non-annualised SPC against days between scans showing no evidence of strong non-linear relationships and relatively stable thinning across timepoints (eFigure 3). Furthermore, uncorrected data analysed with LMMs (eTable 3) and leave-one-site-out analyses (eTable 4) also produced similar findings.

### Cortical thinning and neuroreceptor maps

Cortical thickness SPC was negatively correlated with 5-HT_1A_ (rho = -0.470, P_spin_ = 0.001), 5-HT_2A_ (rho = -0.321, P_spin_ = 0.009) and 5-HT_4_ (rho = -0.620, P_spin_ <.001) receptor topology, indicating greater thinning in receptor-rich regions (Figure 3). Correlations with the remaining maps were non-significant (rho = -0.163 to 0.122, eTable 6, eFigure 4).

**Figure 3.**
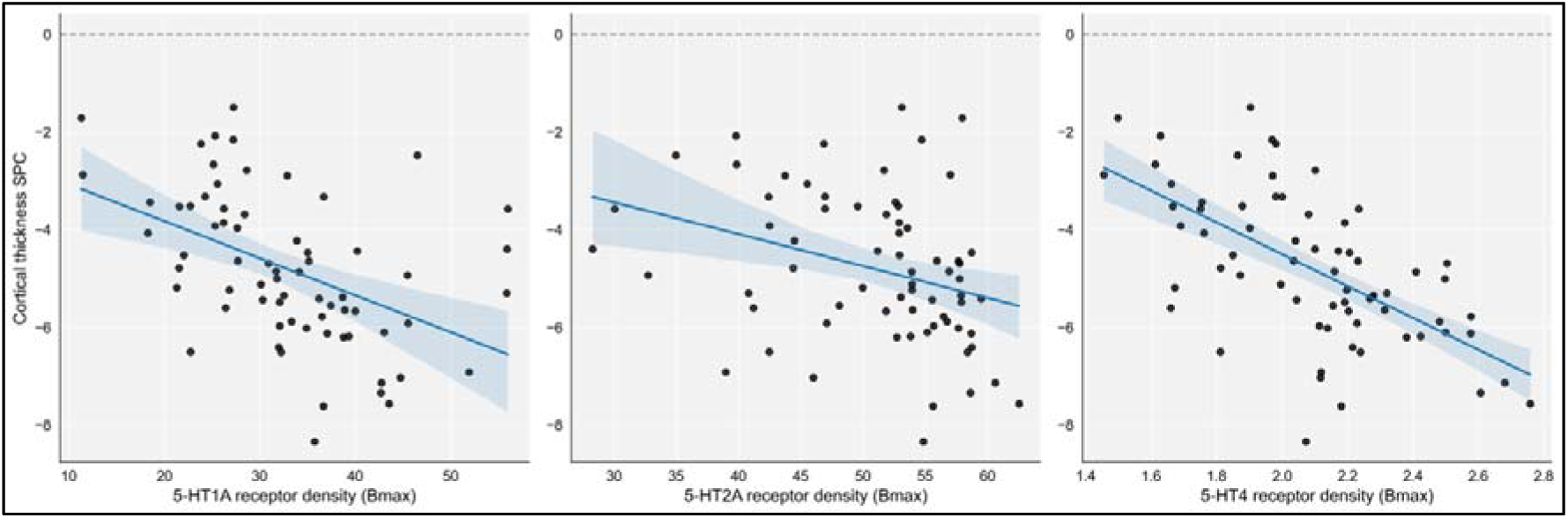
Correlations between SPC of cortical thickness and receptor density maps for 5-HT_1A,_ 5-HT_2A,_ 5-HT_4_. P-values for the correlations, accounting for spatial autocorrelation, are reported in the main text. The shaded area represents the confidence interval.

### Relationships between structural changes and clinical variables

Subcortical volume SPCs were not significantly associated with the percentage change in total or positive symptom severity during clozapine treatment (eTables 7-8). Prior to FDR correction, cortical thickness SPC in the right caudal anterior cingulate cortex (Estimate (E) = 0.61, P = 0.036), right caudal middle frontal cortex (E = 0.60, P = 0.013), right post-central gyrus (E = 0.74, P = 0.040), right superior frontal cortex (E = 0.62, P = 0.026) and bilateral rostral middle frontal cortex (Left: E = 0.74, P = 0.012, Right: E = 0.94, P = 0.007) were positively associated with percentage change in total symptom severity (eTable 7), such that greater thinning was associated with greater symptom improvement. Associations between cortical thickness SPC in the right caudal anterior cingulate cortex (P = 0.022), right post-central gyrus (P = 0.018) and bilateral rostral middle frontal cortex (Left: P = 0.007, Right: P = 0.007) and total symptom severity remained significant after removal of potential outliers. Similar patterns were observed when analyses were restricted to clozapine naïve patients (eTable 9-10). However, none of these associations survived FDR correction. There were no significant associations between cortical thinning and percentage change in positive symptom severity (eTable 8).

Prior to FDR correction, both positive and negative associations between clozapine dose or plasma level and subcortical volume SPC or cortical thickness SPC were detected (eTables 11-12), however none survived FDR correction.

### Relationships between baseline structural features and subsequent changes in symptom severity

There were no significant associations between subcortical volumes, cortical thickness or LGI prior to starting clozapine and the subsequent change in total or positive symptom severity following clozapine treatment (eTables 13-16).

## Discussion

Using a large, combined dataset of 143 individuals with TRS, we examined longitudinal changes in brain structure during clozapine treatment and their relationships with underlying normative cortical features and clinical response. In line with our hypotheses, volumetric reductions in the amygdala, hippocampus, thalamus, caudate, putamen and nucleus accumbens, enlargement of the lateral ventricles and widespread cortical thinning occurred during clozapine treatment. Our analysis additionally revealed an increase in pallidal volume during clozapine treatment. The spatial pattern of cortical thinning was associated with serotonin receptor distributions (5-HT_1A_, 5-HT_2A_ and 5-HT_4_), indicating that regions with higher normative receptor density showed greater thinning during clozapine treatment. Contrary to our hypotheses, after FDR correction neither structural features prior to initiating clozapine nor their change over time were associated with improvement in symptom severity during clozapine treatment.

Previous studies in smaller cohorts (two of which (25,27) are included the current dataset) have reported longitudinal volume reductions in the caudate (24–27), putamen (25), thalamus and hippocampus (27) as well as enlargement of the lateral ventricles (25,27) during clozapine treatment. Our study combines five international datasets to create a much larger cohort and adds to the previous findings to also show pallidal volume increases. Whilst previous studies have not reported increases in pallidal volumes during clozapine treatment, the pallidum is consistently reported as enlarged in schizophrenia relative to healthy controls (6), and may increase with illness duration (7), which would be consistent with our TRS sample. We additionally observed widespread cortical thinning during clozapine treatment, extending previous findings of predominantly frontal and temporal cortical thinning (25,28,29). Interestingly secondary analysis indicated the magnitude of change in subcortical volume and cortical thickness was relatively stable across the range of follow-up intervals, which would be consistent with early alterations rather than progressive trajectories.

Participants in our study had received treatment with non-clozapine antipsychotics for an average of 10 years prior to clozapine initiation. As treatment with non-clozapine antipsychotics is also associated with reductions in subcortical volumes and cortical thickness (15–21), this suggests that the reductions in subcortical volume and cortical thickness that we observed over ∼5 months of clozapine treatment may be additional to structural alterations that had occurred in the preceding years. Brain structural changes may reflect mechanisms related to withdrawing from other antipsychotic medications during switching to clozapine, mechanisms related to starting clozapine, or a combination. It is also possible that brain structural changes could relate to the peripheral metabolic effects of clozapine (78–80). Treatment with non-clozapine antipsychotics has been associated with increases in caudate and putamen volumes (81,82), so the decreases observed here may reflect reversal of earlier medication-related increases. While it is possible that the observed structural changes may also reflect on-going illness mechanisms (11,83), the magnitude of change observed over the clozapine treatment period when patients had already been unwell for many years suggests that switching antipsychotic medication to clozapine results in further reductions in subcortical volume and cortical thickness.

Cortical thinning during clozapine treatment showed negative relationships with cortical 5-HT_1A_, 5-HT_2A_ and 5-HT_4_ receptor distributions, indicating that regions with higher normative serotonergic receptor density exhibited greater thinning. Clozapine has high-affinity inverse agonist / antagonist activity at 5-HT_2A_ receptors (*K_i_* 5.4 nM), and moderate-affinity agonist activity at 5-HT_1A_ receptors (*K_i_* 120 nM) (30,84) but does not have documented affinity at 5-HT_4_. Serotonergic signalling plays an important role in cortical plasticity and reorganisation (45) and may influence cortical thickness. For example, selective serotonin re-uptake inhibitors have been associated with increases in cortical thickness in major depressive disorder (85), and PET studies in bipolar disorder show an inverse relationship between 5-HT_1A_ binding and cortical thickness (86). Psilocybin, a potent 5-HT_2A_ agonist, produces functional reorganisation in human neuroimaging studies (87) and increases synaptogenesis and spine density in animal models (88,89), suggesting that regions with high serotonergic receptor density may be particularly neuroplastic. Our findings partially align with Tuominen et al. (43) who also reported a negative association between antipsychotic-related cortical thinning and 5-HT_2A_ and 5-HT_4_ receptor distributions, but not 5-HT_1A_. This suggests serotonergic topological associations with cortical thinning may be shared across antipsychotics rather than being clozapine-specific, consistent with the affinity of some other antipsychotics at 5-HT_2A_ and 5-HT_1A_ receptors. It also suggests the observed pattern is unlikely to reflect effects of withdrawal or reductions of other antipsychotics. While clozapine has been shown to influence microglial activation and inflammatory signalling (90,91), we found no association with TSPO, likely reflecting the tracer’s limited cellular specificity. Similar to Tuominen et al., (2025), we also found no associations between clozapine-related cortical thinning and receptor maps indexing dopamine, choline, glutamate or GABA systems, suggesting that these neurotransmitters do not relate to regional vulnerability to antipsychotic-related cortical thinning. However, direct longitudinal comparisons of cortical thinning during treatment with clozapine compared to other antipsychotics would be required to determine similarities and differences.

As expected, symptom severity improved during the clozapine observation period, with a numerical improvement in 89% of participants. However, structural alterations were largely unrelated to changes in total or positive symptom severity. Previous reports have linked changes in caudate (26), thalamus and putamen volumes (27) and cortical thickness (28) with symptom improvement during clozapine treatment, but these associations were not observed here. Although we observed a weak relationship between cortical thinning and symptom change, these associations did not survive FDR correction. It is possible that structural changes relate to specific symptom domains not assessed here, for example, depression and anxiety scores, which have been inversely associated with cortical thickness in schizophrenia and bipolar disorder (92). Alternatively, brain structural changes may occur in parallel with symptom improvement during clozapine treatment but are unrelated. Baseline measures of brain structure and gyrification did not predict clinical response, suggesting limited prognostic value. These measures instead may be more informative earlier in illness (61,93,94). The lack of association with brain structural measures highlights the need to explore alternative predictors of clozapine response, such as functional neuroimaging (95), brain glutamate (35) or perfusion (40,96).

### Strengths and limitations

This study integrated data from five international cohorts spanning the UK, Ireland, the US, Chile and South Korea, harmonised using a common processing pipeline. The sample is ∼4 times larger than previous longitudinal studies of brain structural changes during clozapine treatment (25,27,28,49,56), improving statistical power, generalisability and representation. However, as data were integrated retrospectively, protocol differences existed between cohorts, including variation in prior clozapine exposure, although excluding participants previously exposed to clozapine did not meaningfully alter our results. Data on changes in other antipsychotic prescriptions, including dose reductions or discontinuation during clozapine initiation, were not available and may have influenced the observed structural changes. In addition, the absence of a comparator group treated with a non-clozapine antipsychotic limits our ability to disentangle medication effects from illness-related progression, or to attribute the observed changes specifically to clozapine. Although we observed some evidence of associations between structural changes and clozapine dose at follow-up, these did not survive FDR correction, and effect directions were inconsistent. Future studies could better characterise concomitant antipsychotic exposure during clozapine initiation or compare patients with TRS initiating clozapine to those switching to another antipsychotic, to clarify whether observed effects are clozapine specific. However, as clozapine is the only recommended antipsychotic for TRS, the latter may present some practical and ethical challenges. Finally, normative PET maps are based on healthy controls and represent a pre-morbid receptor density, which may not accurately reflect receptor distributions in individuals with TRS.

### Conclusion

In conclusion, clozapine initiation is associated with widespread brain structural changes, including subcortical volume reductions, pallidal and ventricular enlargement and widespread cortical thinning. The spatial pattern of cortical thinning may align with regions of high normative serotonergic receptor density, suggesting a potential role for serotonergic mechanisms in cortical remodelling following clozapine initiation. Brain structural features prior to clozapine initiation and their changes during clozapine treatment appear largely unrelated to improvements in total or positive symptoms.

## Supporting information

Supplementary_eTables

## Data Availability

All data produced in the present study are available upon reasonable request to the authors

## Acknowledgements

The authors would like to thank the following individuals for their contributions to data collection and study coordination: Annie Blazer, K.N. Roy Chengappa and Charles E. Kahn (Pittsburgh); Javiera Vasquez, Juan Aguirre, Camila Diaz Dellarossa, Daniella Barbagelata, Juan Ramirez-Mahaluf, Maria Jose Alfaro, Patricio Carvajal-Paredes and Ruben Nachar (Santiago); and Inkyung Park and Sun Young Moon (Seoul).

