## Supplementary_materials for "Longitudinal brain structural changes during clozapine treatment: associations with neuroreceptor architecture and clinical response"

**eAppendix 1.** Site Inclusion/Exclusion Criteria……………………………………………… pp. 2-3
**eFigure 4.** Associations between cortical thickness SPC and receptor density profiles…….... p.8
**References**……………………………………………………………………………… pp. 9-10

**eAppendix 1.** Site Inclusion/Exclusion Criteria

| Site/Study | Inclusion Criteria | Exclusion criteria |
|---|---|---|
| London | • ICD-10 diagnosis of schizophrenia or schizoaffective disorder<br>• Treatment-resistant illness<br>• About to start clozapine titration as part of routine care<br>Evidence of TRS:<br>• ≥2 previous trials of non-clozapine antipsychotics<br>• Each within recommended dose range<br>• Each for at least 6 weeks | • Pregnancy<br>• MRI contraindications<br>• Drug dependency (DSM-IV)<br>• Clozapine prescribed within the previous 3 months |
| Galway | • Treatment-resistant schizophrenia<br>• About to start clozapine titration as part of routine care<br>Evidence of TRS:<br>• Failure to respond to ≥2 antipsychotics<br>• At least one atypical antipsychotic trial<br>• Persistent moderate–severe positive and/or negative symptoms | • Any previous clozapine trial<br>• Learning disability<br>• History of neurological illness or head injury<br>• Treatment with oral steroid within previous 3 months<br>• Alcohol / drug dependency (DSM-IV) |
| Pittsburgh | • Age 18–60<br>• Diagnosis of schizophrenia or schizoaffective disorder<br>Evidence of TRS:<br>• BPRS score ≥4 (moderate) on:<br>• Hallucinatory behaviour<br>• Unusual thought content<br>• Conceptual disorganization<br>• ≥2 failed non-clozapine antipsychotic trials<br>• Each lasting at least 6 weeks | • No previous adequate clozapine trial (≥4 weeks)<br>• Substance-induced psychotic disorder<br>• Concurrent electroconvulsive therapy<br>• Neurological or medical conditions affecting brain function<br>• Significant suicidal or homicidal risk<br>• MRI contraindications |

| | | |
|---|---|---|
| Santiago | - Age 16-50<br>- Lifetime history of psychotic disorder (ICD-10 schizophrenia or schizoaffective)<br>- ≥2 failed non-clozapine antipsychotic trials<br>- Each lasting at least 6 weeks at recommended doses<br>- Clinician initiating clozapine | - Neurological or medical conditions affecting brain function or structure.<br>- Drug or alcohol dependence.<br>- MRI contraindications.<br>- No previous exposure to clozapine |
| Seoul | • Age ≥19<br>• DSM-5 schizophrenia spectrum or other psychotic disorder<br>• Treatment-resistant symptoms<br>• About to start clozapine titration in standard care<br>• Clozapine-naive<br>Evidence of TRS:<br>• ≥2 non-clozapine antipsychotic trials<br>• Within recommended dosage<br>• Each ≥6 weeks<br>• Referred for clozapine initiation | • Medication with significant interaction with clozapine within 2 weeks<br>• Alcohol or substance use disorder (DSM-5)<br>• MRI contraindications<br>• Pregnancy or intention to conceive |

**eFigure 1.** Residual baseline cortical thickness (averaged across regions) before and after ComBat harmonisation.

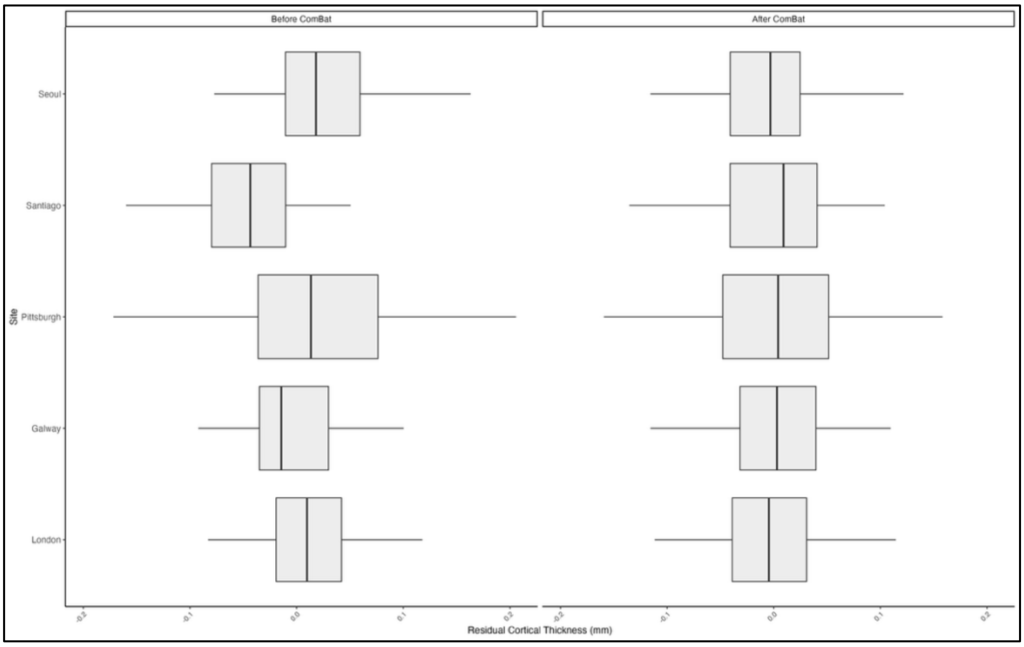

*Note*: ComBat harmonisation was implemented using the neuroHarmonize package, with age, sex, and illness duration included as covariates. Age and illness duration were modelled using generalized additive model (GAM) terms to account for potential non-linear effects. The ComBat model was trained on baseline data and subsequently applied to follow-up data prior to calculation of annualised symmetrised percentage change (SPC) for each cortical region. Analyses were conducted across 68 cortical regions defined by the Desikan-Killiany atlas. For visualisation purposes only, cortical thickness values were additionally averaged across regions to aid interpretability of the ComBat-adjusted data.

**eAppendix 2.** PET maps

| Map | Tracer | Measure | N Participants | Age Mean (SD) | Reference |
|---|---|---|---|---|---|
| D1 | [$^{11}$C]SCH23390 | BPnd | 13 | 33 (13) | Kaller et al., 2017 (1) |
| D2 | [$^{11}$C]FLB-457 | BPnd | 55 | 32.45 (9.69) | Sandiego et al., 2015 (2) |
| 5-HT1A | [11C]CUMI-101 | Bmax | 8 | 28.40 (8.8) | Beliveau et al., 2017 (3) |
| 5-HT1B | [11C]AZ10419369 | Bmax | 36 | 27.80 (6.9) | Beliveau et al., 2017 (3) |
| 5-HT2A | [11C]CIMBI-36 | Bmax | 29 | 22.6 (2.7) | Beliveau et al., 2017 (3) |
| 5-HT4 | [11C]SB207145 | Bmax | 59 | 25.9 (5.3) | Beliveau et al., 2017 (3) |
| 5-HTT | [11C]DASB | Bmax | 100 | 25.1 (5.8) | Beliveau et al., 2017 (3) |
| 5-HT6 | [11C]GSK215083 | BPnd | 30 | 36.6 (9.04) | Radhakrishnan et al., 2018; 2020 (4,5) |
| M1 | [11C]LSN3172176 | BPnd | 24 | 40.45 (11.71) | Naganawa et al., 2021 (6) |
| α4β2 | [18F]Flubatine | $V_T$ | 30 | 33.50 (10.71) | Hillmer et al., 2016 (7) |
| NMDA | [18F]GE-179 | $V_T$ | 29 | 40.9 (12.7) | Hansen et al., 2022 (8) |
| mGluR5 | [11C]ABP688 | BPnd | 73 | 19.9 (30.04) | Smart et al., 2019 (9) |
| GABA | [11C]Flumazenil | Bmax | 16 | 26.6 (8.0) | Nørgaard et al., 2021 (10) |
| CBF | | ASL | 922 | range 8-22 | Satterthwaite et al., 2014 (11) |
| Glucose | [18F]FDG | CMRglc | 33 | 25.4 (2.6) | Vaishnavi et al., 2010 (12) |
| Oxygen | [18F]FDG | CMRO2 | 33 | 25.4 (2.6) | Vaishnavi et al., 2010 (12) |
| HDAC | [11C]Martinostat | SUVR | 8 | 28.6 (7.6) | Wey et al., 2016 (13) |
| TSPO | [11C]PBR28 | SUVR | 6 | 57.8 (8.1) | Lois et al., 2018 (14) |
| SV2A | 11C-UCB-J | BPnd | 76 | 48.9 (18.4) | Finnema et al., 2018; Naganawa et al., 2021(6,15) |

**eFigure 2.** Non-annualised subcortical volumes SPC and days between scans.

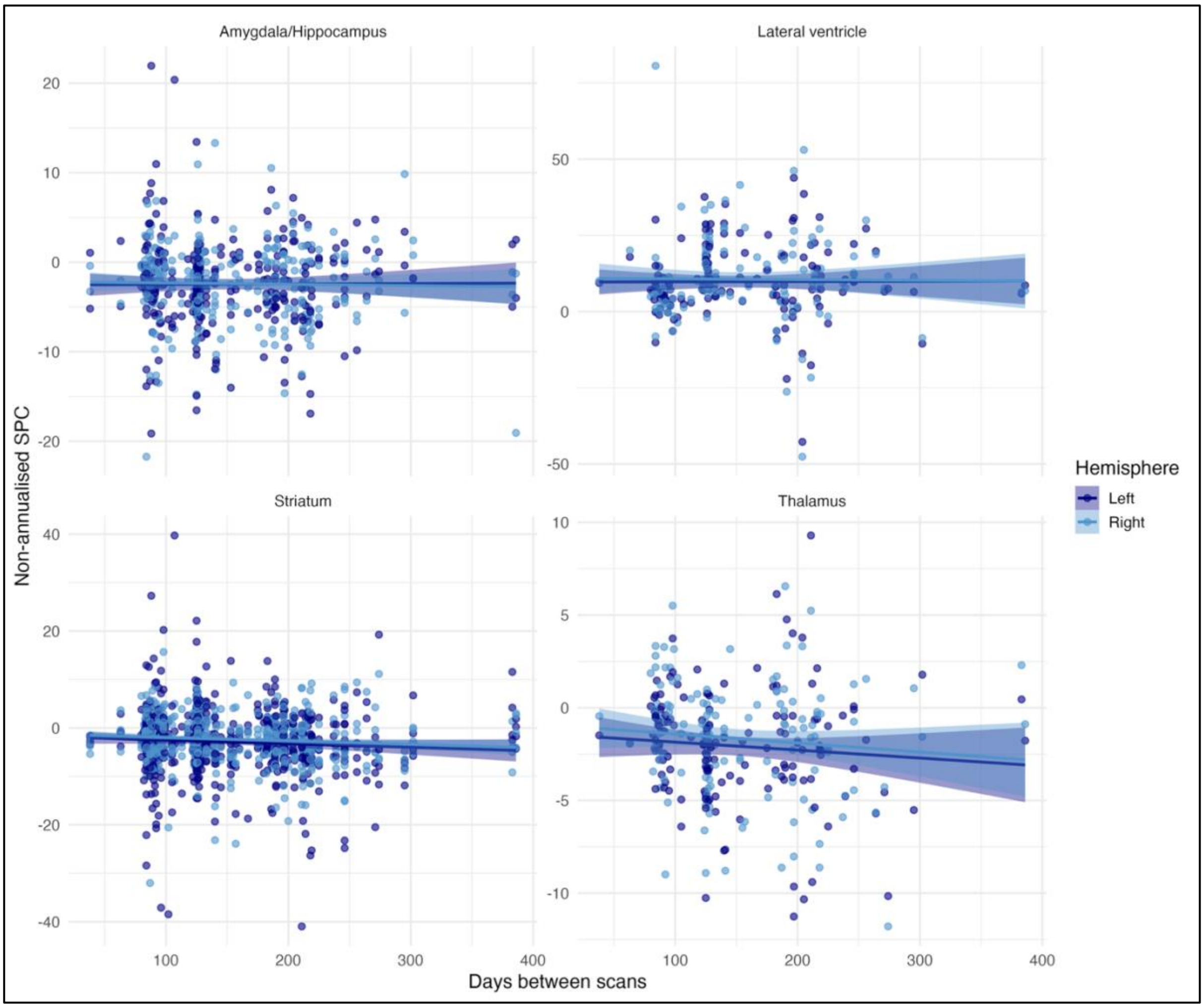

*Note*: Analyses were conducted on 16 bilateral subcortical regions (left and right hemispheres analysed separately). For visualisation purposes only, regional values were averaged within anatomically related groups (striatal regions and amygdala-hippocampus) to assess linearity. These aggregations were not used in statistical analyses. The figure presents linear regression fits with 95% confidence intervals.

**eFigure 3.** Non-annualised cortical thickness SPC and days between scans.

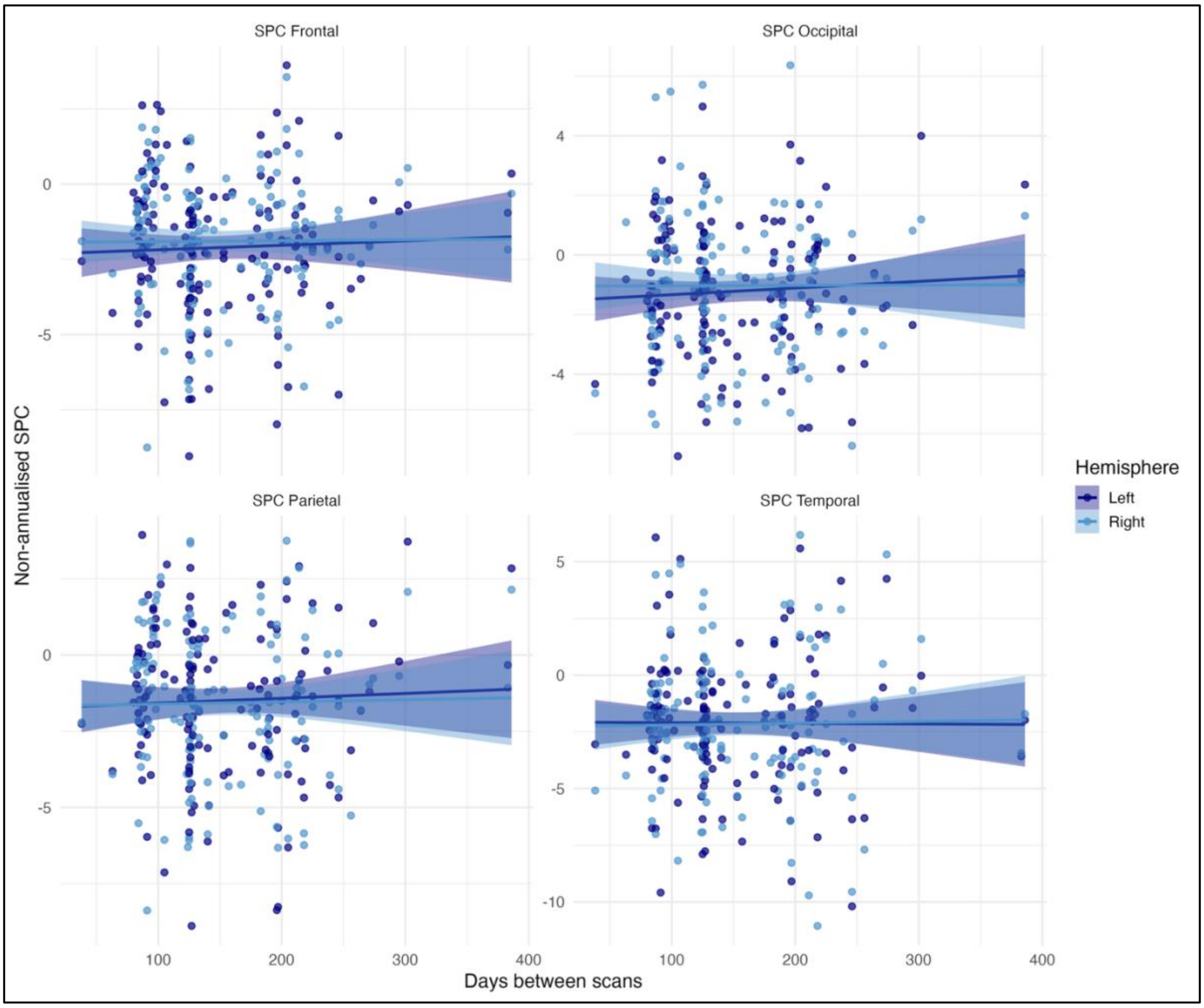

*Note*: Cortical thickness values were derived from the Desikan-Killiany atlas. Analyses were conducted on 68 cortical regions, with left and right hemispheres analysed separately. For visualisation purposes only, regional values were averaged across major cortical lobes (frontal, occipital, parietal and temporal) to assess linearity. These aggregations were not used in statistical analyses. The figure presents linear regression fits with 95% confidence intervals.

**eFigure 4.** Associations between cortical thickness SPC and receptor density profiles.

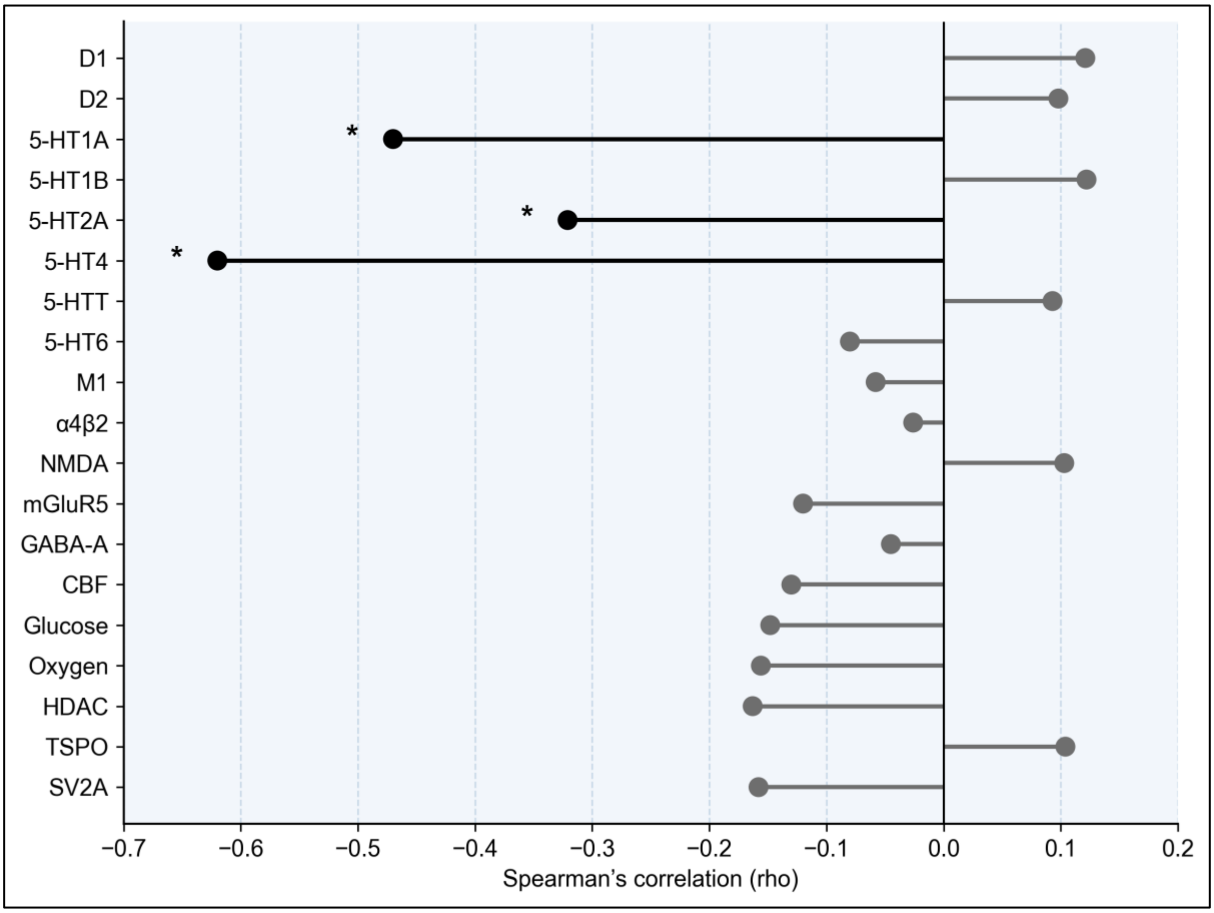

*Note*: * Denotes significance after spatial-autocorrelation correction ($P_{spin}$<0.05).
